# Network-based analyses identify GFAP as a cross-domain hub linking synaptic, neuronal, and inflammatory markers in Alzheimer’s disease

**DOI:** 10.64898/2026.05.22.26353857

**Authors:** Chiara Trasciatti, Andrea Pilotto, Chiara Tolassi, Flavio Ragni, Elena Marcello, Monica Moroni, Stefano Bovo, Caterina Martinuzzo, Silvia Pelucchi, Salvatore Caratozzolo, Irene Girotto, Laura D’Andrea, Ramona Stringhi, Andrea L. Benedet, Ilaria Pola, Henrik Zetterberg, Nicholas Ashton, Giuseppe Jurman, Monica di Luca, Alessandro Padovani

## Abstract

Alzheimer’s disease (AD) is characterized by complex alterations in synaptic, glial, neuronal and inflammatory markers. Given its emerging role at the interface of synaptic dysfunction and inflammation, the astrocytic marker GFAP may represent a cross-domain hub linking synaptic, neuronal and inflammatory alterations. Using multivariate and network-based analyses we examined the relationships among cerebrospinal fluid (CSF) biomarkers of astrocytic activation and synaptic failure, inflammation, and neurodegeneration in biologically confirmed AD patients and healthy controls (HC).

We studied 60 AD patients and 40 HC. CSF concentrations of Neurogranin, SNAP-25, CAP2, NfL, GFAP, IL-1α, IL-1β, IL-8, MCP-1, TNF-α were measured. Associations were assessed using Spearman correlations, LASSO regression, and network analysis to characterize multivariate dependency structures.

Compared with controls, AD patients showed significantly higher CSF levels of Neurogranin, SNAP-25, CAP2, NfL, GFAP, IL-1β, TNF-α. In AD, synaptic biomarkers were strongly intercorrelated and associated with astroglial activation, inflammatory markers, and tau-related pathology. Network analysis identified GFAP as a cross-domain hub linking synaptic, inflammatory, and neurodegenerative domains in AD. In controls, GFAP was mainly associated with neuronal injury markers.

Network-based modelling revealed a disease-related reorganization of biomarker connectivity in AD, with GFAP occupying a central cross-domain position, supporting a systems-level view of AD pathophysiology.

## INTRODUCTION

In recent years, substantial progress has been made in defining the biological architecture of Alzheimer’s disease (AD). Biomarkers reflecting amyloid-β deposition, tau pathology, and neurodegeneration are now embedded within contemporary diagnostic criteria and have substantially improved the biological characterization of the disease [1,2]. Nevertheless, the pathophysiology of AD extends beyond these canonical domains and includes additional processes that may emerge early and contribute directly to disease expression, progression, and heterogeneity [3–5].

Among these processes, synaptic dysfunction has gained increasing attention as a core feature of AD [6]. Experimental, imaging, and neuropathological studies indicate that synaptic loss is an early and progressive event, closely linked to cognitive decline and only partially captured by conventional markers of neuronal injury [7].

Among CSF biomarkers of synaptic injury, Neurogranin and SNAP-25 (Synaptosomal-Associated Protein 25) are the most extensively investigated and are consistently elevated in individuals with AD compared with neurologically healthy controls. Increased concentrations of these markers have been associated with poorer cognitive performance and downstream neurodegeneration [8– 15]. In addition, CAP2 (Cyclase-Associated Protein 2), a postsynaptic actin-binding protein involved in dendritic spine remodeling during synaptic plasticity events, has emerged as a novel marker of synaptic failure, showing increased CSF concentrations in AD and strong associations with tau pathology [16–18].

At the same time, growing evidence suggests that synaptic dysfunction in AD is embedded within a broader biological context involving glial activation and inflammatory signaling. Astrocytes and other glial cells are increasingly recognized as active modulators of synaptic integrity, amyloid-related processes, and neuronal vulnerability rather than passive responders to tissue damage [19,20]. In this framework, glial fibrillary acidic protein (GFAP), a marker of astrocytic activation, has emerged as a particularly relevant candidate for capturing a biological interface between amyloid-related pathology, synaptic failure, and downstream neurodegenerative processes [14,21–25]. However, the extent to which GFAP relates to synaptic and inflammatory biomarker profiles in vivo remains insufficiently defined. Importantly, available biomarkers of synaptic injury, astrocytic activation, inflammation, and neuronal damage appear to co-vary only partially, suggesting overlapping but non-identical biological pathways [26].

These observations indicate that the relationships among synaptic, glial, inflammatory, and neurodegenerative markers in AD are unlikely to be fully described by isolated pairwise associations or by simple linear models. Rather, they are more plausibly embedded within an interdependent biological system in which the relevance of each biomarker depends on its position within the overall network of interactions [27,28]. For this reason, network-based approaches may provide a useful framework to investigate how these biomarker domains are organized in relation to one another [29] and whether specific markers occupy a cross-domain role within the AD biomarker architecture [30,31].

Accordingly, in the present study we applied complementary multivariate and network-based analyses to age- and sex-adjusted CSF biomarkers in a biologically confirmed cohort of patients with AD and healthy controls, with the aim of characterizing the relationships, between astrocytic activation and synaptic dysfunction, inflammation, and neurodegeneration in vivo.

## METHODS

### Study Population

The study population consisted of individuals who were assessed for cognitive disturbances at the outpatient Neurodegenerative Clinic of the Brescia University Hospital, Italy, along with age-matched healthy control subjects. This cross-sectional study included consecutive patients diagnosed with MCI or mild dementia who underwent CSF assessments. All patients underwent routine blood analyses, magnetic resonance imaging (MRI), and a standardized clinical assessment including Mini-Mental State Examination (MMSE) and the Clinical Dementia Rating Scale (CDR).

Exclusion criteria included: (1) cognitive deficits or dementia not fitting clinical criteria for AD, DLB, or FTD; (2) significant cortical or subcortical infarcts or abnormal brain iron accumulation on imaging; (3) other neurological disorders or medical conditions potentially contributing to cognitive deficits; (4) psychiatric conditions such as bipolar disorder, schizophrenia, or history of drug or alcohol abuse; (5) recent traumatic events or acute fever/inflammation that could affect

CSF and plasma biomarkers. The diagnosis of AD was carried out clinically and confirmed biologically according to CSF AD-pattern Aβ-42/p-Tau 181 ratio > 11.1[1,32,33]. For biomarkers comparison, a cohort of non-neurological controls (HC, n=40) who underwent CSF analyses for acute headache but normal MRI, standard CSF analyses and NfL were included [34]. The study was approved by the local ethics committee (DMA study, NP 1471 and Neuromultibio study, NP 5285, Brescia Ethics Committee) and followed the Helsinki Declaration, with informed consent obtained from all participants.

### CSF collection and analysis

At enrollment, 3 milliliters of CSF from each participant were collected. Lumbar puncture was performed under fasting conditions according to the standardized protocol of the outpatient clinic, from 09:00 to 11:00 in the morning, after clinical informed written consent was obtained. CSF was collected in sterile polypropylene tubes and gently mixed to avoid gradient effects. CSF was centrifuged and first processed for standard biochemical analyses, whereas two milliliters of CSF were stored in cryotubes at –80°C before biomarker testing. Only patients with normal routine measures were included in further analyses.

CSF Aβ-42 (Amyloid beta 1–42), Aβ-40 (Amyloid beta 1–40), p-Tau 181 (Phosphorylated Tau at threonine 181), and t-tau (Total Tau) were measured using the Lumipulse G assays (Fujirebio) on the LUMIPULSE G600II for diagnostic standard analyses. All CSF sample analyses for inflammatory, neuronal and glial markers were conducted by researchers who were blind to clinical status, at the Laboratory of Advanced Biological Markers at the University of Brescia, utilizing the Neurology 2-Plex Advantage Kits for NfL (Neurofilament light chain), GFAP and SNAP-25 kit (single-plex) on the SIMOA SR-X platform from Quanterix, Billerica, MA. CSF cytokine concentrations including interleukin IL-8 (Interleukin-8), IL-1β (Interleukin-1 beta), IL-1α (Interleukin-1 alpha), MCP-1 (Monocyte Chemoattractant Protein-1), TNF-α (Tumor Necrosis Factor alpha) and Neurogranin were measured using an Invitrogen ProcartaPlex 10-plex assay on the Luminex MagPix platform. For SIMOA and Luminex analyses, all biomarkers were measured in duplicates with CV < 20% and the mean of the technical duplicate assessments was used for final analyses.

CAP2 was measured using an enzyme-linked immunosorbent assay (ELISA) performed at the Department of Pharmacological and Biomolecular Sciences (University of Milan), as previously reported[17]. Briefly, the CSF samples were diluted at 1:20, and their CAP2 concentration was determined using a commercially available ELISA (catalog number IK5163; Immunological Sciences, Rome, Italy). This assay has high sensitivity and specificity for CAP2 detection; no significant cross-reactivity or interference between CAP2 and its analogs was observed. The mean of duplicate assessment was used for final analyses.

### Statistical analyses

Normality was assessed using the Shapiro-Wilk test and Q-Q plots. To compare demographic and clinical characteristics and CSF markers between AD and HC, categorical variables were analyzed using the χ^2^ (Chi-square) test, and continuous variables using Student’s t-test, Welch’s t-test and Mann-Whitney test, as appropriate. Spearman’s correlations, adjusted for age and sex, were classified as weak (ρ=0.10–0.30), moderate (ρ=0.31–0.50), or strong (ρ ≥ 0.51). Differences between Spearman correlation coefficients across groups were compared using Fisher’s r-to-z transformation. This transformation converts correlation coefficients into a normally distributed variable while preserving the original sign of the correlation, enabling direct statistical comparisons. The resulting z-scores were used for statistical comparisons. A z-test was performed, and a p < 0.05 threshold was applied to the z-score matrices to identify significantly altered connections in patients compared to HC.

To identify biomarkers associated with synaptic marker levels, LASSO regression, a regularization technique that performs variable selection and reduces overfitting, was applied separately to the AD and HC groups [35]. All variables were standardized prior to analysis to ensure comparability and proper penalization across different measurement scales. The regularization parameter (λ) was tuned using 5-fold cross-validation to balance model complexity and prediction accuracy, and 3- and 4-fold cross-validation were also run as robustness checks. Variables with coefficients reduced to zero by LASSO were excluded, while those with non-zero coefficients were retained as significant predictors. Model performance was assessed using the in-sample coefficient of determination (R^2^) to evaluate goodness of fit. Collinearity diagnostics were assessed using the Variance Inflation Factor (VIF < 5), ensuring absence of problematic multicollinearity among predictors prior to regularization.

To better understand the complex relationships between variables, potentially overlooked by classical univariate or predictive modeling approaches, network analysis was implemented to visualize overall biomarker connectivity. Network analysis was conducted on biomarker residuals adjusted for age and sex. Association matrices were estimated using a rank-based normalized (Gaussian-rank) transformation of the residuals followed by correlation, corresponding to a nonparanormal approach that relaxes distributional assumptions while preserving monotonic dependencies [36]. Network visualization employed a force-directed layout based on the Fruchterman-Reingold algorithm [37], implemented in the qgraph framework for network visualization [38], in which node positions result from a global optimization of attractive and repulsive forces, emphasizing overall network topology. Networks were thresholded by statistical significance using false-discovery-rate (FDR) control [39]. Edge width encoded the magnitude and sign of correlations. For descriptive purposes, node-level centrality was quantified using strength, defined as the sum of the edge weights incident to each node [40]. For visualization purposes, node size was scaled according to node degree (number of edges connected to each biomarker), providing a visual indication of local connectivity within the network.

All inflammatory biomarkers were included to preserve the system-level structure of the network and avoid selection based on univariate significance. Additional analyses were performed restricting the network to cytokines significantly differing between groups.

All analyses were conducted using R Statistical Software (https://www.r-project.org/) or Python version 3.11.13 (https://www.python.org/). Statistical significance was defined at α=0.05, and all tests were two-tailed.

## RESULTS

The study assessed an extensive panel of CSF biomarkers in 60 AD patients (median age 76 years, interquartile range, IQR: 70.00-79.00 years) and 40 age and sex-matched neurologically healthy controls (HC). Demographic and clinical characteristics and CSF biomarker levels are shown in Table 1.

**Table 1.**
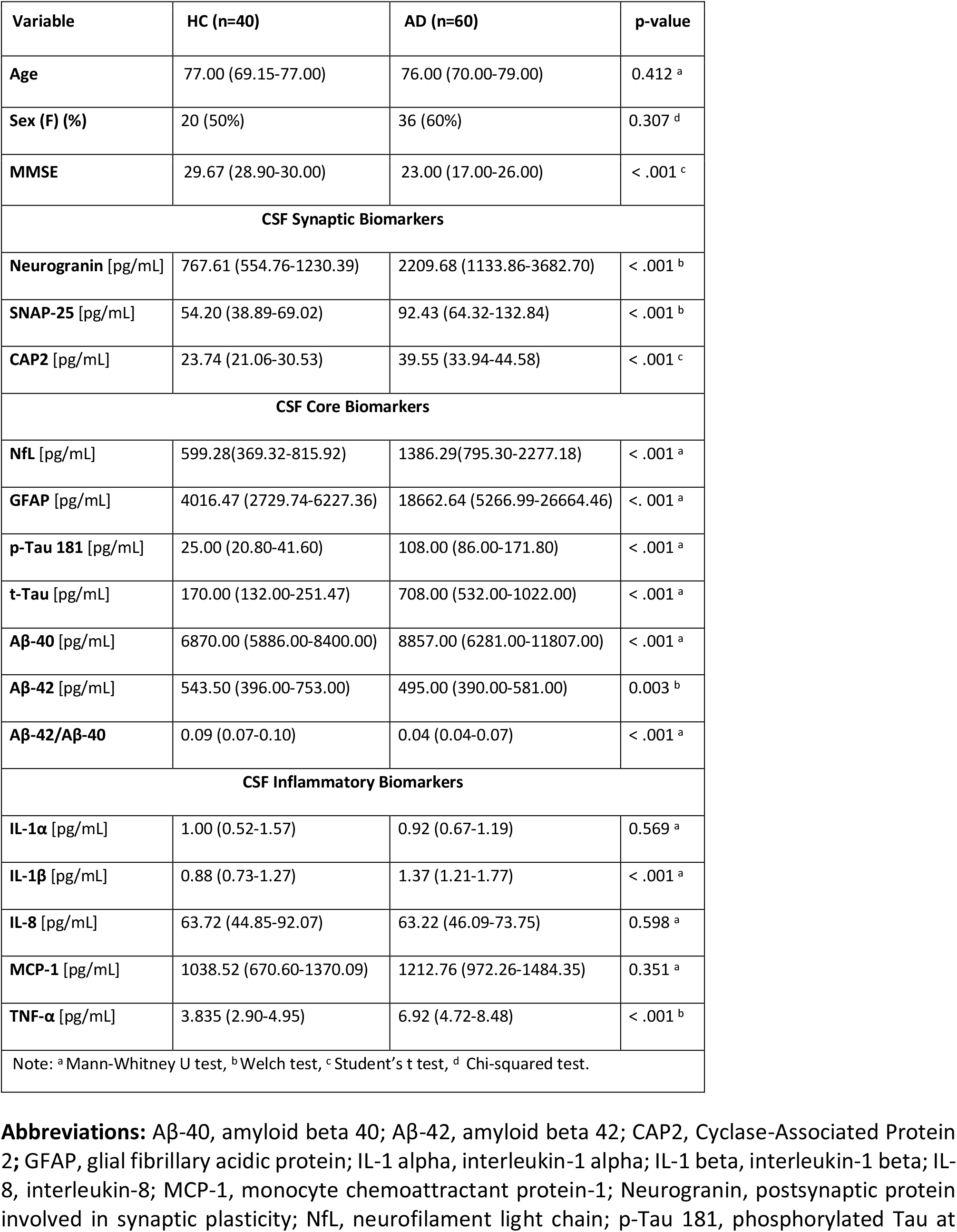
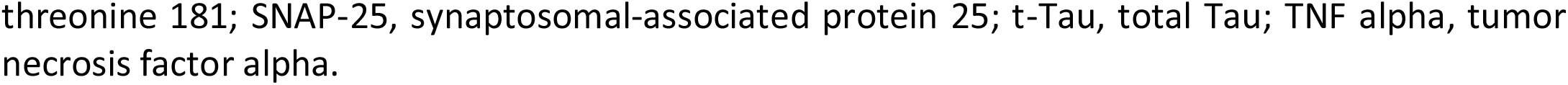
Demographics, clinical and CSF biomarkers characteristics in healthy controls and AD patients. Continuous variables are reported as median and interquartile range (25th–75th percentile) and categorical variables are reported as numbers and percentages (n, %).

| Variable | HC (n=40) | AD (n=60) | p-value |
| --- | --- | --- | --- |
| Age | 77.00 (69.15-77.00) | 76.00 (70.00-79.00) | 0.412 <sup>a</sup> |
| Sex (F) (%) | 20 (50%) | 36 (60%) | 0.307 <sup>d</sup> |
| MMSE | 29.67 (28.90-30.00) | 23.00 (17.00-26.00) | < .001 <sup>c</sup> |
| **CSF Synaptic Biomarkers** |  |  |  |
| Neurogranin [pg/mL] | 767.61 (554.76-1230.39) | 2209.68 (1133.86-3682.70) | < .001 <sup>b</sup> |
| SNAP-25 [pg/mL] | 54.20 (38.89-69.02) | 92.43 (64.32-132.84) | < .001 <sup>b</sup> |
| CAP2 [pg/mL] | 23.74 (21.06-30.53) | 39.55 (33.94-44.58) | < .001 <sup>c</sup> |
| **CSF Core Biomarkers** |  |  |  |
| NfL [pg/mL] | 599.28(369.32-815.92) | 1386.29(795.30-2277.18) | < .001 <sup>a</sup> |
| GFAP [pg/mL] | 4016.47 (2729.74-6227.36) | 18662.64 (5266.99-26664.46) | < .001 <sup>a</sup> |
| p-Tau 181 [pg/mL] | 25.00 (20.80-41.60) | 108.00 (86.00-171.80) | < .001 <sup>a</sup> |
| t-Tau [pg/mL] | 170.00 (132.00-251.47) | 708.00 (532.00-1022.00) | < .001 <sup>a</sup> |
| Aβ-40 [pg/mL] | 6870.00 (5886.00-8400.00) | 8857.00 (6281.00-11807.00) | < .001 <sup>a</sup> |
| Aβ-42 [pg/mL] | 543.50 (396.00-753.00) | 495.00 (390.00-581.00) | 0.003 <sup>b</sup> |
| Aβ-42/Aβ-40 | 0.09 (0.07-0.10) | 0.04 (0.04-0.07) | < .001 <sup>a</sup> |
| **CSF Inflammatory Biomarkers** |  |  |  |
| IL-1α [pg/mL] | 1.00 (0.52-1.57) | 0.92 (0.67-1.19) | 0.569 <sup>a</sup> |
| IL-1β [pg/mL] | 0.88 (0.73-1.27) | 1.37 (1.21-1.77) | < .001 <sup>a</sup> |
| IL-8 [pg/mL] | 63.72 (44.85-92.07) | 63.22 (46.09-73.75) | 0.598 <sup>a</sup> |
| MCP-1 [pg/mL] | 1038.52 (670.60-1370.09) | 1212.76 (972.26-1484.35) | 0.351 <sup>a</sup> |
| TNF-α [pg/mL] | 3.835 (2.90-4.95) | 6.92 (4.72-8.48) | < .001 <sup>b</sup> |
| Note: <sup>a</sup> Mann-Whitney U test, <sup>b</sup> Welch test, <sup>c</sup> Student's t test, <sup>d</sup> Chi-squared test. |  |  |  |

**Abbreviations:** Aβ-40, amyloid beta 40; Aβ-42, amyloid beta 42; CAP2, Cyclase-Associated Protein 2; GFAP, glial fibrillary acidic protein; IL-1 alpha, interleukin-1 alpha; IL-1 beta, interleukin-1 beta; IL-8, interleukin-8; MCP-1, monocyte chemoattractant protein-1; Neurogranin, postsynaptic protein involved in synaptic plasticity; NfL, neurofilament light chain; p-Tau 181, phosphorylated Tau at
1 threonine 181; SNAP-25, synaptosomal-associated protein 25; t-Tau, total Tau; TNF alpha, tumor 2 necrosis factor alpha.

AD patients exhibited higher levels of synaptic markers, including Neurogranin, SNAP-25, and CAP2 (p ≤ 0.001), compared to HC (Figure 1, Panel a). They also showed increased NfL, p-Tau 181, t-Tau, and Aβ-40, along with reduced Aβ-42 and a lower Aβ-42/Aβ-40 ratio. Synaptic marker levels did not show any correlation with age, sex or MMSE total score (analysis was separately performed, available as supplementary material).

**Figure 1.**
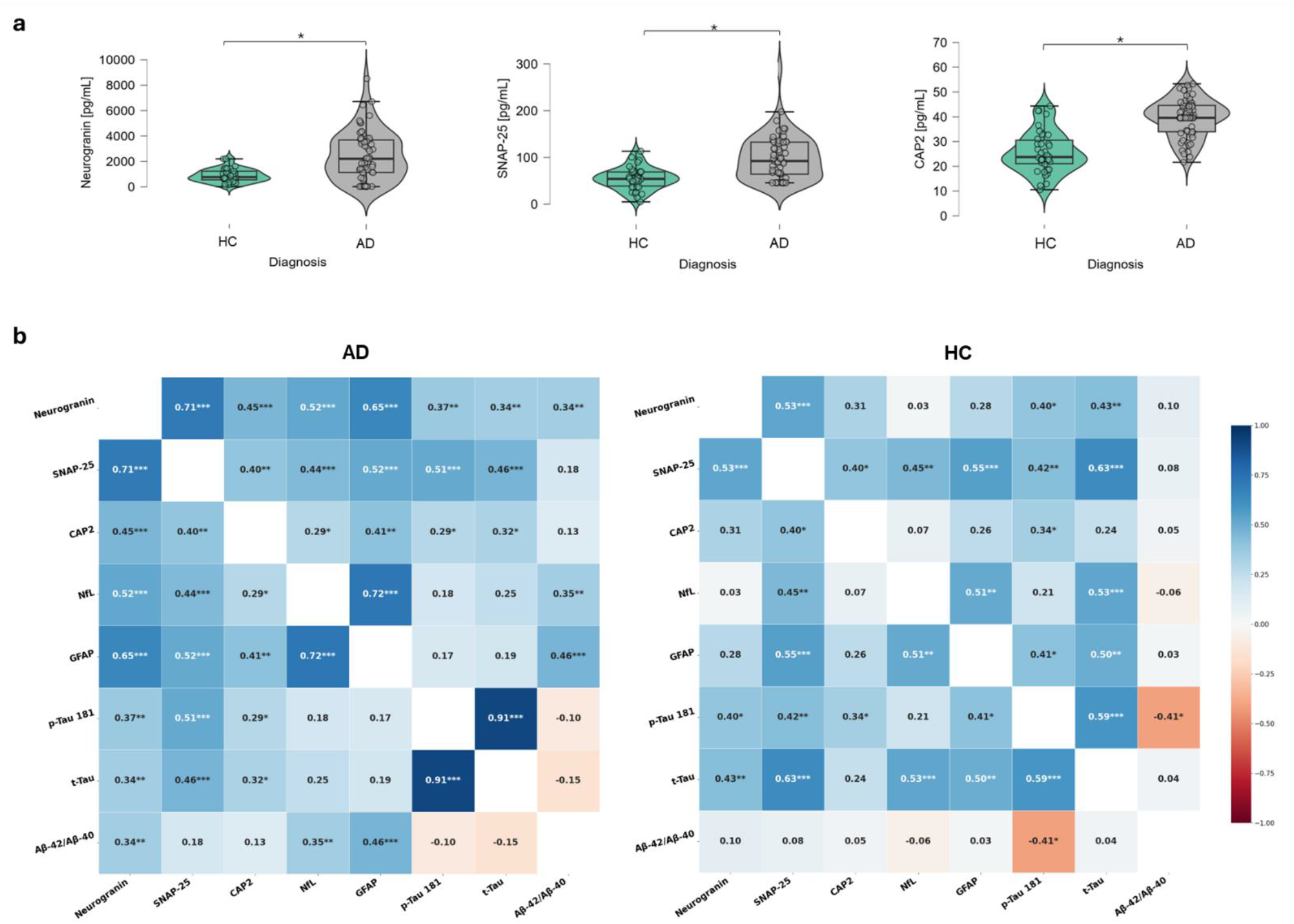
Panel a: Violin plots showing the distribution of synaptic marker concentrations in AD patients and healthy controls, with median and interquartile range indicated. Panel b: Spearman correlation matrix of CSF biomarkers adjusted for age and sex in AD and HC. Color intensity reflects correlation strength and direction (red = negative; blue = positive); asterisks denote statistical significance (* p < 0.05, ** p < 0.01, *** p < 0.001). **Abbreviations:** Aβ-40, amyloid beta 40; Aβ-42, amyloid beta 42; CAP2, Cyclase-Associated Protein 2; GFAP, glial fibrillary acidic protein; Neurogranin, postsynaptic protein involved in synaptic plasticity; NfL, neurofilament light chain; p-Tau 181, phosphorylated Tau at threonine 181; SNAP-25, synaptosomal-associated protein 25; t-Tau, total Tau.

### Correlation Patterns Among Biomarkers

In the correlation analysis, synaptic markers showed distinct patterns of inter-relationship between AD patients and HC. Synaptic markers were strongly inter-correlated in the AD group, with Neurogranin and SNAP-25 showing the strongest positive association (ρ = 0.71, p < 0.001). CAP2 was also positively correlated with Neurogranin (ρ = 0.45, p < 0.001) and SNAP-25 (ρ = 0.40, p < 0.001). In addition, all synaptic markers displayed moderate to strong correlations with GFAP, NfL and tau isoforms (Figure 1b).

In HC, correlations among synaptic markers, as well as between synaptic and other biomarkers, were generally weak and largely non-significant. The correlation matrices for both groups are shown in Figure 1 (Panel b). Moreover, comparison of the two correlation matrices highlighted biomarker pairs with statistically significant differences in their correlation patterns between the two groups (Supplementary Figure 1). Neurogranin exhibited stronger correlations in the AD group with GFAP (Z = 2.29, p = 0.022) and NfL (Z = 2.51, p = 0.012). In addition, p-Tau 181 and t-Tau showed a significantly stronger coupling in AD (Z = 3.85, p < 0.001), while GFAP was more strongly associated with the Aβ-42/Aβ-40 ratio in the same group (Z = 2.19, p = 0.028).

### LASSO Regression Model

To further examine the variables strongly associated with synaptic markers, a LASSO regression approach was applied, separately for CAP2, Neurogranin and SNAP-25. All corresponding coefficients are presented in Table 2.

**Table 2.** LASSO regression coefficients for biomarkers predicting Neurogranin, SNAP-25 and CAP2 in AD and HC. Model fit (R^2^) for each outcome is reported under the column header. Positive coefficients indicate a positive association; negative coefficients indicate an inverse association. Dashes (–) denote predictors excluded from the model.

| Predictor | Neurogranin – AD<br>$R^2 = 0.44$ | Neurogranin – HC<br>$R^2 = 0.36$ | SNAP-25 – AD<br>$R^2 = 0.34$ | SNAP-25 – HC<br>$R^2 = 0.53$ | CAP2 – AD<br>$R^2 = 0.20$ | CAP2 – HC<br>$R^2 = 0.19$ |
| --- | --- | --- | --- | --- | --- | --- |
| NfL | - | – 0.201 | - | 0.100 | - | - |
| GFAP | 0.378 | 0.027 | 0.095 | 0.363 | 0.097 | 0.190 |
| p-Tau 181 | 0.083 | 0.291 | - | - | - | 0.091 |
| t-Tau | - | 0.170 | 0.110 | 0.270 | 0.095 | - |
| Aβ-40 | 0.235 | 0.321 | 0.300 | - | 0.180 | - |
| Aβ-42 | - | - | - | 0.151 | - | 0.031 |

When using Neurogranin as the dependent variable, the model selected GFAP, p-Tau 181 and Aβ-40 in the AD group (R^2^ = 0.44), with the strongest positive contribution from GFAP. In HC group (R^2^ = 0.36), selected predictors were GFAP, NfL, p-Tau 181, t-Tau and Aβ-40 with NfL negatively weighted.

Using SNAP-25 as the dependent variable, the model retained GFAP, t-Tau and Aβ-40 in the AD group (R^2^ = 0.34), all with positive coefficients. In the HC group (R^2^ = 0.53), the selected variables were GFAP, NfL, t-Tau and Aβ-42. GFAP and t-Tau had the highest positive coefficients.

Using CAP2 as the dependent variable, the model retained GFAP, t-Tau and Aβ-40 in the AD group (R^2^ = 0.20), all with positive coefficients. In the HC group (R^2^ = 0.19), the selected variables were GFAP, p-Tau 181, Aβ-42.

Sensitivity analysis using 3- and 4-fold cross-validation showed comparable selection patterns, consistently retaining GFAP among the strongest predictors.

### Network Analysis

In addition to synaptic and neurodegeneration markers, inflammatory biomarkers (IL-1α, IL-1β, IL-8, MCP-1, TNF-α) were integrated in a network analysis to examine their contribution to the overall architecture and their relationship with neuronal, glial and synaptic pathways.

In the AD network, high-weight connections were observed among markers of neuronal injury/neurodegeneration (NfL, t-Tau), astrocyte activation (GFAP), p-tau-related pathology (p-Tau 181), amyloid-related measures (Aβ-40, Aβ-42), and synaptic proteins (Neurogranin, SNAP-25, CAP2), forming a densely interconnected structure. Inflammatory markers (IL-8, IL-1α, TNF-α) displayed multiple cross-domain associations, linking synaptic, glial, and neurodegenerative nodes rather than forming an isolated cluster. Node-level centrality quantified by strength indicated that Neurogranin showed the highest connectivity (strength = 1.58), followed by GFAP (strength = 1.22) and TNF-α (strength = 0.72). GFAP maintained connections spanning synaptic, neurodegenerative, and inflammatory domains, consistent with a cross-domain positioning within the AD network architecture.

The HC network exhibited a more compartmentalized organization. Synaptic, neurodegenerative, and amyloid-related markers formed a core cluster, with Aβ-42 showing multiple direct associations. Inflammatory markers were primarily connected among themselves and showed limited associations with synaptic or neurodegenerative nodes.

Strength-based centrality in the HC network showed that SNAP-25 exhibited the highest connectivity (strength = 1.72), followed by t-Tau (strength = 1.26) and GFAP (strength = 1.07). GFAP showed connections mainly involving neurodegeneration-related markers, without direct inflammatory links.

Network structures are visualized in Figure 2, illustrating the pattern and magnitude of inter-biomarker associations in each group. Restricting the network to inflammatory markers significantly differing between groups (IL-1β, TNF-α) yielded a comparable topology in the AD network, with GFAP maintaining high cross-domain centrality.

**Figure 2.**
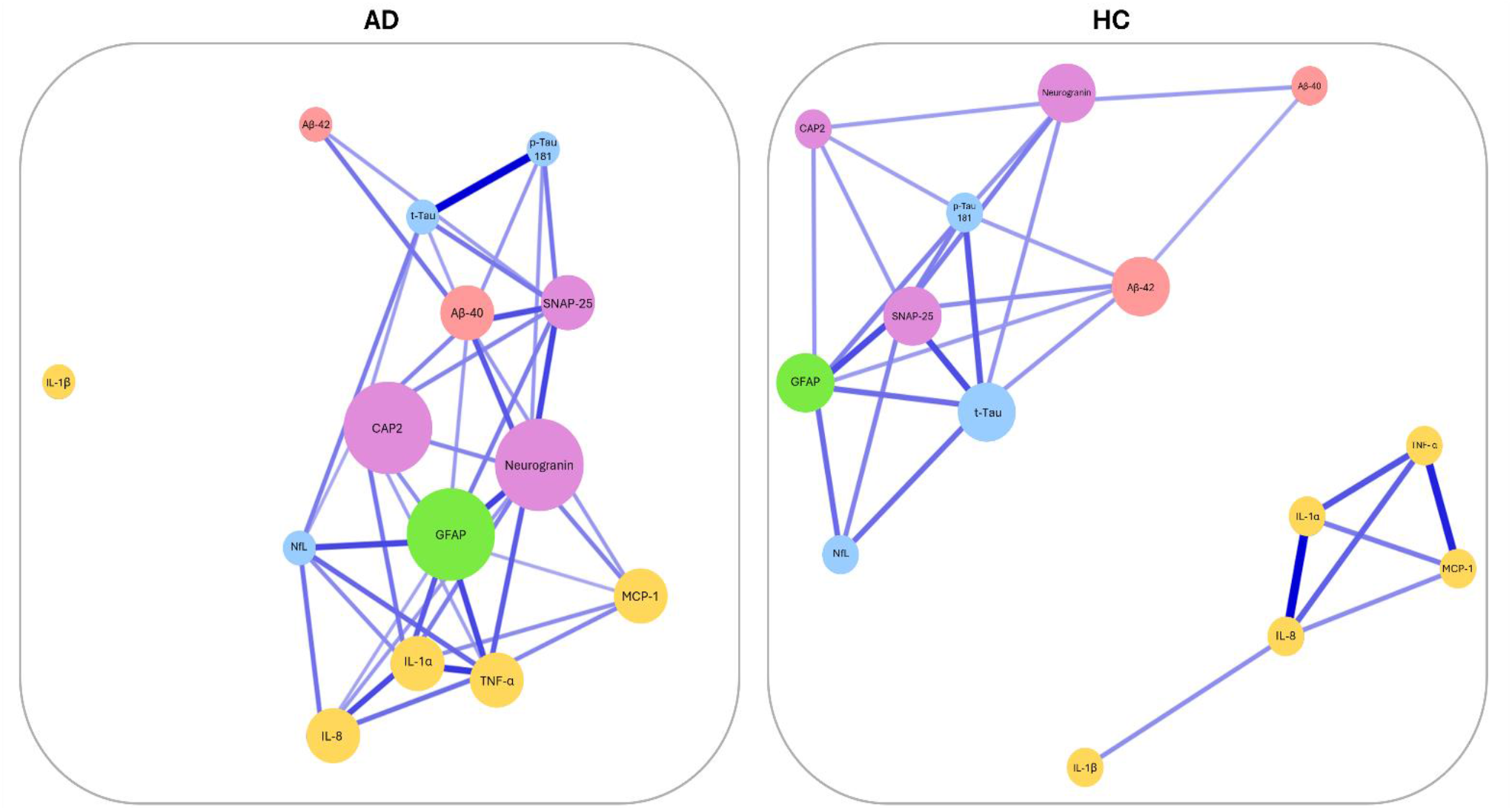
Network analysis of biomarker associations in AD and HC groups. Nodes represent individual biomarkers, colored by functional category: synaptic (purple: Neurogranin, SNAP-25, CAP2); neuronal injury and tau-related neurodegeneration (blue: NfL, p-Tau 181, t-Tau); glial (green: GFAP); amyloid-related (red: Aβ-42, Aβ-40); inflammatory (yellow: IL-1α, IL-1β, IL-8, MCP-1, TNF-α). Node size reflects the number of connections (node degree) for each biomarker in the network. Edge thickness and transparency are proportional to the absolute correlation between biomarker pairs. Abbreviations: AD, Alzheimer’s Disease; HC, healthy controls; Neurogranin; SNAP-25, Synaptosomal-associated protein 25 kDa; CAP2, Cyclase-associated protein 2; NfL, Neurofilament light chain; GFAP, Glial fibrillary acidic protein; p-Tau 181, Phosphorylated tau (Thr181); t-Tau, Total tau; Aβ-40, Amyloid-β 1–40; Aβ-42, Amyloid-β 1–42.

## DISCUSSION

In this study, we investigated the in vivo relationships among CSF biomarkers of astrocytic activation, synaptic dysfunction, neurodegeneration, amyloid pathology, and inflammation in AD using complementary correlation, penalized regression, and network-based approaches. Our findings support the view that synaptic failure in AD is not an isolated process, nor merely a downstream consequence of amyloid or tau pathology [2,41], but rather part of a broader and biologically interconnected system [11,42–44]. Within this multivariate framework, GFAP emerged as a prominent cross-domain marker linking synaptic, neurodegenerative, and inflammatory pathways. These results suggest that astrocytic activation may occupy an integrative position within the biomarker architecture of AD and reinforce the value of network-based approaches for capturing disease-related reorganization across pathological domains, as recently suggested also by the work of Wang and colleagues [7,25,45–48].

Consistent with previous literature, CSF Neurogranin and SNAP-25 concentrations were increased in AD compared with healthy controls [8,12,49]. CAP2, more recently proposed as a synaptic biomarker associated with tau-related processes, was also elevated in AD [16,17]. These observations support the concept that synaptopathy is a core biological component of AD and not simply a late correlation of established neuronal loss. Elevated synaptic markers have also been associated with cognitive decline and disease progression in AD, further reinforcing their relevance to disease expression [50,51].

Beyond these group-level differences, our correlation analyses revealed a disease-specific reorganization of biomarker coupling. In AD, synaptic markers were strongly inter-correlated, with Neurogranin and SNAP-25 showing the strongest association, and CAP2 positively associated with both. This coordinated behavior of presynaptic and postsynaptic markers is consistent with the notion of a biochemical signature of synaptic damage and remodeling in AD and prodromal stages [6]. Importantly, in AD these markers were not confined to a within-domain synaptic pattern but also showed moderate associations with markers of axonal injury, tau pathology, and astrocytic activation [8,16,52]. In particular, the associations of Neurogranin with GFAP and NfL were significantly stronger in AD than in controls, supporting a tighter biological coupling between synaptic dysfunction, astrocyte activation, and neuronal injury in the disease state [41,53–55].

By contrast, in healthy individuals, associations among synaptic and other biomarkers were still detectable, including correlations between synaptic proteins and GFAP and between tau-related measures and synaptic markers [41,46]. However, the overall organization in controls appeared more compartmentalized with correlations tending to remain limited to specific biomarkers pairs rather than extending across multiple pathological domains [55]. This distinction between AD and controls is relevant as it suggests that the disease state is characterized not only by absolute biomarker changes, but also by a restructuring of their interrelationships. In this context, the stronger coupling in AD between GFAP and Neurogranin, as well as with NfL may reflect a closer integration between astrocytic responses and synaptic failure, potentially under conditions driven or amplified by amyloid-related pathology [21,43].

The LASSO regression analyses further supported the close association between glial and synaptic markers. Across models, GFAP was consistently retained as a predictor of synaptic proteins in AD. This recurrent selection pattern is noteworthy because it was observed across different synaptic outcomes and remained stable in sensitivity analyses using alternative cross-validation strategies. At the same time, the moderate proportion of explained variance indicates that additive linear approaches capture only part of the underlying biological complexity. This point is important, as it justifies the complementary use of network-based analyses to describe higher-order interdependencies that are not readily captured by conventional regression models [35].

The network analyses provided the clearest evidence that AD is associated with a cross-domain reorganization of biomarker connectivity [56]. In the AD network, synaptic markers were embedded within a densely interconnected structure linking glial activation, neurodegeneration, tau pathology, amyloid-related measures, and inflammatory markers. This pattern is consistent with the view that AD pathophysiology reflects system-level reorganization rather than isolated biomarker alterations [27,28,31]. Within this framework, GFAP showed a prominent topological position, maintaining connections across synaptic, neurodegenerative, and inflammatory domains. Although Neurogranin displayed the highest overall connectivity strength, GFAP was distinctive in its cross-domain embedding, suggesting that astrocyte activation may act as an interface among multiple pathological processes rather than simply mirroring one biological axis.

This interpretation is biologically supported. Astrocytes are increasingly recognized as active regulators of synaptic homeostasis, metabolic support, inflammatory signaling, and responses to amyloid pathology. In particular, astrocytic processes are integral to synapses, not merely passive separators of the synaptic cleft. The traditional view of the synapse as purely pre- and postsynaptic neuronal elements has evolved: astrocytes express glutamate receptors, respond to neurotransmitters, and release gliotransmitters, actively participating as partners in the tripartite synapse [57]. Accordingly, increased GFAP concentrations may reflect more than a nonspecific reaction to injury; they may index a state in which astrocytic activation is tightly coupled with synaptic vulnerability and with broader disease-related remodeling of the neural microenvironment [19,20,22–24].

In parallel, inflammatory markers such as IL-8, IL-1α, and TNF-α were not segregated into an isolated inflammatory cluster but were integrated into the broader AD network. This observation supports the view that inflammatory mediators may participate in the same pathological configuration linking glial activation, synaptic dysfunction, and neuronal injury, rather than representing a fully independent process [20,58].

Importantly, these network configurations should not be interpreted as evidence of direct causality. Rather, they reflect patterns of statistical interdependence within a multivariate biological system. Nevertheless, from a systems biology perspective, such cross-domain embedding is informative because it identifies biomarkers that occupy structurally relevant positions within the overall architecture of disease-related interactions [29,30]. In this respect, GFAP appears less relevant as an isolated marker level than as a biomarker whose relational profile changes substantially in AD.

In contrast, the network observed in HC displayed a more compartmentalized organization. Inflammatory markers were largely confined to intra-inflammatory associations, and cross-domain connections between synaptic, glial, and neurodegenerative markers were less extensive. Within this configuration, GFAP maintained primarily neurodegeneration-related associations but did not exhibit the same degree of cross-domain integration observed in AD. This difference in overall architecture supports the interpretation that AD is characterized by a reorganization of biomarker interrelationships, with stronger integration between glial activation, inflammation, and synaptic dysfunction.

Overall, these findings suggest that the biological relevance of synaptic biomarkers in AD may be better understood when considered within a wider network of interacting pathological processes. From this perspective, the study adds to growing evidence that glia-synapse interactions are integral to AD pathophysiology and may help explain part of the heterogeneity observed across patients [14,59]. In fact, these results support the use of network-informed approaches to identify biologically meaningful patterns that extend beyond simple group comparisons or pairwise associations.

This study is not without limitations. First, the cross-sectional design and the relatively modest sample size preclude causal inference and limit conclusions regarding the temporal evolution of biomarker interactions. Longitudinal studies will be required to determine whether the observed network configurations precede clinical progression, track disease stages, or identify biologically distinct subtypes. Second, the biomarker panel, although multidomain, remains incomplete, and the inclusion of additional synaptic, microglial, astroglial, and neurodegenerative markers could further refine the characterization of these complex biological networks. Third, the control group, while neurologically healthy, was not drawn from a population-based sample, which may affect generalizability. Finally, network topology and centrality measures should be interpreted cautiously in datasets of this size and ideally be confirmed in larger independent cohorts.

Despite these limitations, our findings highlight the potential value of network-based analytical frameworks for studying AD as a systems-level disorder. Future studies should examine whether glia-related connectivity patterns are associated with clinical trajectories, cognitive resilience, or therapeutic response, and whether such approaches can inform biomarker-guided stratification.

In conclusion, our results indicate that synaptic dysfunction in AD is embedded within a complex multivariate network linking glial, neurodegenerative, amyloid, and inflammation. Within this architecture, GFAP emerges as a biologically relevant cross-domain marker, reinforcing the importance of glia-synapse interactions and the utility of network-based approaches for capturing structured dependency patterns in AD.

## Data Availability statement

The dataset is not publicly available due to privacy restrictions but is available from the corresponding author on reasonable request.

## Acknowledgments

The study has been supported by the European Union -Next Generation EU - NRRP M6C2 - Investment 2.1 Enhancement and strengthening of biomedical research in the NHS-PNRR – PNRR PNRR-Health PNRR-MAD-2022-12376110.

The authors wish to thank the study participants who took part in this research. We are very grateful for the assistance received from the Neurology department of the Spedali Civili of Brescia.

## Author contributions

CTr, Api and APa contributed to the conception and design of the study. CTr and APi performed the data analysis and prepared the figures. CTo, IG, EM and SP contributed to the biological analyses. CM, AR and SC contributed to data acquisition. FR, MM, SB, EM, SP, LDA, RS, ALB, IP, HZ, NJA, GJ, MdL and APa contributed to drafting and revising the manuscript.

