## Supplementary Figure 1 for "Network-based analyses identify GFAP as a cross-domain hub linking synaptic, neuronal, and inflammatory markers in Alzheimer’s disease"

**Supplementary Figure 1.** *Comparison of pairwise Spearman correlations between biomarkers in the AD and HC cohorts after adjustment for age and sex. The heatmap displays Z-scores representing the difference between group-specific correlation coefficients. Only statistically significant differences (p < 0.05, two-tailed) are shown. Positive values indicate stronger correlations in the AD group relative to HC (blue), and negative values indicate stronger correlations in HC (red).*

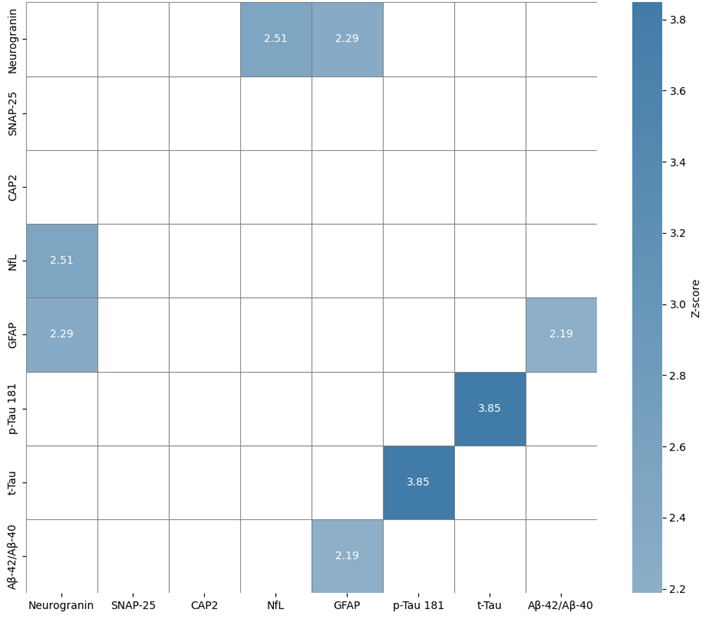

Abbreviations: Neurogranin; SNAP-25, Synaptosomal-associated protein 25 kDa; CAP2, Cyclase-associated protein 2; NfL, Neurofilament light chain; GFAP, Glial fibrillary acidic protein; p-Tau 181, Phosphorylated tau (Thr181); t-Tau, Total tau; Aβ-40, Amyloid-β 1–40; Aβ-42, Amyloid-β 1–42.
